# Atmospheric PM2.5 before and after Lockdown in relation to COVID-19 Evolution and daily Viral Counts: Could Viral Natural Selection have occurred due to changes in the Airborne Pollutant PM2.5 acting as a Vector for SARS-CoV-2?

**DOI:** 10.1101/2021.01.10.21249548

**Authors:** Yves Muscat Baron

## Abstract

**Background:** Genes coding for SARS-CoV-2 have been detected on the microscopic airborne pollutant particulate matter, which has been suggested as a vector for COVID-19 transmission. Lockdown in China has been shown to be associated with significant reduction in pollution including the particulate matter component which coincided with the appearance of a viral mutant (Clade G) which steadily displaced the original Clade D after lockdown. The reason why Clade G developed a fitness advantage is as yet unknown. This paper examines the possible role of airborne particulate matter PM2.5 as selective pressure determining viral Clade predominance and further shedding light on the mode of SARS-CoV-2 transmission.

**Methods:** The average levels of PM2.5 of a number of cities were obtained from the Air Quality Index (AQI), a real-time assessment of atmospheric pollution. The daily average PM2.5 levels were assessed between January 23rd and April 29^th^ 2020 determined by the timeline when viral counts in Beijing and other cities were available. Daily viral counts of Clades D and G were available starting from the 12^th^ February as determined by the scientific literature published in August 2020. The cities chosen were Beijing, Sheffield, Nottingham, Sydney and Cambridge because of their substantially elevated viral counts compared to other cities. Cities as opposed to vaster areas/nations were chosen as PM2.5 levels vary across regions and countries.

**Results:** For the time period assessed, the Beijing PM2.5 pattern initiated with highly elevated mean PM2.5 levels of 155.8µg/m3 (SD+/-73.6) during high viral counts, followed by 82.1µg/m3 (SD+/-44.9) (p<0.04) when the viral counts decreased. In all the other cities assessed, the pattern differed whereby the PM2.5 levels increased significantly over the preceding baseline contemporaneously with the viral count rise. The changes in these cities’ PM2.5 levels were on average 31.5µg/m3 before viral counts rose and 56.35µg/m3 contemporaneous with viral count rise. The average levels of PM2.5 in these cities started to decrease one week after lockdown to 46µg/m3 when measured over 2 weeks post-lockdown.

As regards the viral counts from data retrieved from Beijing, the latter part of the bell-shaped curve and a subsequent smaller curve of the viral count was available for evaluation. The average viral count for Clade D in Beijing was 11.1(SD+/-13.5) followed by a mean viral count for Clade G was 13.8(SD+/-9.2). Conversely in all the other cities besides Beijing, the viral counts averaged 45.8 for Clade D and 161 for Clade G. The variation in viral counts between cities suggests the strong possibility of variation in the availability of sampling between cities.

The newer variant, Clade G demonstrated viral counts initially appearing in mid-February in Beijing to later displace Clade D as the dominant viral Clade. The appearance of Clade G coincided with the decreasing gradient of PM2.5 levels. A number of significant correlations were obtained between PM2.5 levels and the viral count in all the cities reviewed.

**Conclusion:** COVID-19 viral counts appear to increase concomitant with increasing PM2.5 levels. Viral counts of both Clades correlated differentially with PM2.5 levels in all the cities assessed. The significantly highly elevated PM2.5 levels in Beijing resulted in correlating mainly with Clade D, however Clade G began to appear with decreasing PM2.5 levels, suggesting the beginnings for the initial SARS-CoV-2 Clade evolution. Clade G, the newer variant was able to flourish at lower levels of PM2.5 than Clade D. Clade G may possibly have utilized other sources of particulate matter as a viral vector, such as that derived from tobacco smoking, whereby 66% of Chinese males are smokers and 70% of the Chinese non-smoking population are exposed to 2^nd^ hand smoking.

## INTRODUCTION

COVID-19 infection has reappeared in most countries, the recrudescence delineated as the “2nd wave”. This increase has been attributed to the reversal of certain measures endorsing social distancing which was legally enforced in most countries affected. Concomitant with the increase in COVID-19 infection incidence, the previous reduction in atmospheric pollution following national and regional lockdowns has been shown to have been partially reversed. This reversal also involved a recrudescence of elevated levels of the pollutant particulate matter PM2.5.

In the United States, a nationwide study by Wu et al 2020 indicated a link between long-term exposure to particulate matter PM2.5 and COVID-19 related mortality.^1^ Genes coding for SARS-CoV-2 have been found to be adherent to particulate matter.^2^ A study of a large number of Chinese cities demonstrated a 2% increase in COVID-19 new cases, with every 10μg/m3 increase in atmospheric PM2.5.^3^ A paper by Comunian et al (2020) suggested that through two avenues, particulate matter may encourage COVID-19 infection and transmission.^4^ The first avenue suggests that the deleterious effects of particulate matter on pulmonary microbial defences encourage COVID-19 to colonize and infect the respiratory system. The second suggests that particulate matter may actually act as a vector for transmission of COVID-19 infection.

This paper examines the possibility that pollution changes catalyzed COVID-19 infection and also its evolution, strengthening the hypothesis that particulate matter may act as a vector and co-factor for the Coronavirus Pandemic.

## METHODS

The average levels of particulate matter PM2.5 of a number of cities were obtained from the Air Quality Index. The Air Quality Index is a real-time measurement of atmospheric pollutants, including PM2.5.^5^ The daily average PM2.5 levels were assessed between January 23rd and April 29^th^ 2020 determined by the timeline when viral counts in Beijing occurred. The 23^rd^ January 2020 was chosen as a starting point when the lockdown was implemented in the Hubei Province of which Wuhan is the principal city. April 29^th^ was the day when the viral count in all cities chosen had decreased significantly.

Daily viral counts of Clades D and G were obtained from a paper by Korber et al. ^6^ This paper obtained viral samples from a large number of centres across the globe in varying numbers. The cities chosen were Cambridge (U.K.), Sheffield (U.K.), Nottingham (U.K.), Sydney (Australia) and Beijing (Temple of Heaven) because of their substantially elevated viral counts compared to other cities. The substantial number of viral counts reduced the error from sampling bias. Cities as opposed to vaster areas were chosen as particulate matter PM2.5 levels vary across regions and countries.

## RESULTS

The daily viral counts of Clades D and G obtained from the paper by Korber et al indicated a pattern peculiar for Beijing as opposed to the other four cities.^6^ This paper obtained viral samples from a large number of centres across the globe in varying numbers. The cities chosen were Beijing, Sheffield, Nottingham, Sydney and Cambridge because of their substantially elevated viral counts compared to other cities so as to reduce the error from sampling bias.

In Beijing, Clade D initially predominated until the 4^th^ March when Clade G started to appear. Viral counts began to appear in the British cities and in Sydney in appreciable amounts after the 4^th^ of March. Following the 4^th^ of March, Clade G became the predominant viral Clade to gradually displace Clade D not only in Beijing but in all the cities assessed.

Similar to the viral counts, the measurements of particulate matter PM2.5 in Beijing showed a different pattern compared to the other cities assessed. In Beijing the PM2.5 pattern initiated with highly elevated mean PM2.5 levels of 155.8µg/m3 (SD+/-73.6) during elevated viral counts. When the viral counts in Beijing decreased, the PM2.5 also decreased by a margin of 36% to 82.1µg/m3 (SD+/-44.9) (p<0.04).

In the British cities and in Sydney, the pattern differed from that of Beijing, whereby the PM2.5 levels increased significantly over the preceding baseline concomitant with viral count rise. The changes in city PM2.5 levels were as follows with the first figure representing the PM2.5 levels before viral counts rose and the second number demonstrating the PM2.5 levels contemporaneous with viral count rise: Sheffield 26.9µg/m3 (SD+/-5.9) and 60.3µg/m3 (SD+/-33.3)(p<0.00048); Nottingham 30.3µg/m3 (SD+/-7.5) and 60.3µg/m3 (SD+/-33.3)(p<0.028), Sydney 33.4µg/m3 (SD+/-7.3) and 51µg/m3 (SD+/-27)(p<0.04), and Cambridge 35.4µg/m3 (SD+/-11.1) and 53.8µg/m3 (SD+/-21.5)(p<0.005).

The levels of PM2.5 in these cities altered after regional and national lockdown. The levels of PM2.5 in these cities decreased significantly one week after regional and national lockdown. The PM2.5 levels one week after were Sheffield 46.6µg/m3(SD+/-27.3), Nottingham 44.2µg/m3 (SD+/-15), Sydney 45µg/m3 (SD+/− 20), Cambridge 49µg/m3 (SD+/-22.7) when measured for 2 weeks. The results of the PM2.5 levels are depicted in Table 1.

**Table 1.** Particulate Matter PM2.5 before Viral Count rise, PM2.5 during Viral Count Rise and PM2.5 one week after Lockdown.

| **Table 1.** | PM2.5 before Viral Count rise | PM2.5 during Viral Count Rise | PM2.5 one week after Lockdown |
| --- | --- | --- | --- |
| Beijing | 155.8µg/m<sup>3</sup> (SD+/-73.6) | 82.1µg/m<sup>3</sup> (SD+/-44.9) | 80.1µg/m<sup>3</sup> (SD+/-44.5) |
| Sheffield | 26.9µg/m<sup>3</sup> (SD+/-5.9) | 60.3µg/m<sup>3</sup> (SD+/-33.3) | 46.6µg/m<sup>3</sup>(SD+/-27.3) |
| Nottingham | 30.3µg/m<sup>3</sup> (SD+/-7.5) | 60.3µg/m<sup>3</sup> (SD+/-33.3) | 44.2µg/m<sup>3</sup> (SD+/-15) |
| Sydney | 33.4µg/m<sup>3</sup> (SD+/-7.3) | 51µg/m<sup>3</sup> (SD+/-27) | 45µg/m<sup>3</sup> (SD+/-20) |
| Cambridge | 35.4µg/m<sup>3</sup> (SD+/-11.1) | 53.8µg/m<sup>3</sup> (SD+/-21.5) | 49µg/m<sup>3</sup> (SD+/-22.7) |

Viral counts varied between the five cities included in this study. As regards the viral counts from data retrieved from Beijing, the latter part of the bell-shaped curve and a subsequent smaller curve of the viral count was available for evaluation. The average viral count for Clade D in Beijing was 11.1(SD+/-13.5) followed by a mean viral count for Clade G was 13.8(SD+/-9.2). Conversely the initial full bell-shaped curves of viral counts were available for the cities of Sheffield, Nottingham, Sydney and Cambridge. The respective viral counts for these cities were Sheffield Clade D 20.9(SD+/-21.7) and Clade G 71.2(SD+/-44.6), Nottingham Clade D 31.6(SD+/-18.3) and Clade G 81.8(SD+/-42.5), Sydney Clade D 31.6(SD+/-21.1) and Clade G 51.8(SD+/-43.3), Cambridge Clade D 99.1(SD+/-64.3) and Clade G 439(SD+/-359). The variation in viral counts between cities suggests the strong possibility of variation in the availability of sampling between cities.^6^

A number of significant correlations were obtained between PM2.5 levels and the viral count. Some correlations were obtained for both Clades for the cities assessed and some for only one viral Clade. PM2.5 in Beijing correlated with Clade D viral counts (R=0.68 p<0.001) but not for Clade G. Both viral Clades of Sheffield correlated with PM2.5: Clade D (R=0.73 p<0.0003), Clade G (R=0.74 p<0.004). Similarly both viral Clades of Nottingham correlated with PM2.5: Clade D (R=0.45 p<0.02), Clade G (R=0.43 p<0.04). For Sydney and Cambridge only Clade G correlated with PM2.5 Sydney (R=0.53 p<0.04) and Cambridge (R=0.79 p<0.0002). For Cambridge the correlation between PM2.5 and Clade D just missed statistical significance (R=0.53 p<0.0505).

Similar significant correlations were obtained when the PM2.5 was compared with viral counts assessed a week later. This exercise was done to allow for the viral incubation period to take root. In the case of Cambridge the correlation between PM2.5 and the Clade G viral count one week later was significant (R=0.62 p<0.002).

The data was analysed for normality and all the data were found to be nonparametric. The Mann Whitney U test was applied for comparing nonparametric variables of both groups of cities and the Spearman Rank test was applied for nonparametric correlations.

## DISCUSSION

### Mutational Potential of COVID-19 and PM2.5

Earlier this year the genome of COVID-19 was sequenced and was found to consist of a single RNA strand containing approximately 30,000 purine and pyrimidine bases. Following the first sequence more than 100,000 SARS-CoV-2 genomes have been sampled indicating that over 13,000 mutations have occurred over a period of nine months.^7^

The rate of genomic modification for SARS-CoV-2 has been estimated to occur around twice a month, significantly lower than that of Influenza virus.^7^ Through these mutations, there exists the possibility of natural selection occurring as an adaptation to the environmental milieu, including the atmospheric levels of particulate matter. This may be of singular importance if particulate matter is proven to be a vector for COVID-19, especially if the particulate matter implicated originates from tobacco smoking.^8,9^

A prominent mutation that has dominated the COVID-19 genomic landscape involves the viral spike protein gene at D614G position. The D614G mutation involves a single genomic alteration with the replacement of aspartic acid by glycine at the amino acid D614 position of the spike protein. In vitro, the D614G mutation has demonstrated increased cellular infectivity, however this does not seem to be consistently the case in vivo.^7^

The modification in the Clade genome appears to result in an alteration in the phenotype of the spike protein peptide configuration. The original Clade D has its spike protein’s three peptides aligned in a “closed” configuration whereas the Clade G spike protein peptides are consistently found in an “open” orientation.^10^ COVID-19 viral adherence to the respiratory goblet cells depends on spike protein’s trimeric peptide orientation. The spike protein adheres to respiratory goblet cells’ angiotensin II receptors if at least two of its three peptides are positioned in an “open orientation”.^11^ As a corollary the “open” orientation may also affect SARS-CoV-2 adherence to particulate matter if the latter acts as its vector.

Clade G’s different trimeric peptide configuration may reflect significant changes to the functional components of the spike protein receptor binding domain. Molecular dynamic computer simulations have demonstrated a complex network of salt bridges, hydrophobic sites, hydrogen bonding and electrostatic interactions between the receptor-binding domain of the COVID-19 Spike protein and the angiotensin II receptor.^12^ Consequently, the salt and water content of the viral environs, including a potential vector, may be relevant to the viral pathogenic behaviour.

### PM2.5 as a Viral Vector and Evolution of COVID-19

This review indicated that prior to the viral count increase, the PM2.5 level was significantly low in all the cities except Beijing. As regards Beijing, only the latter half of the bell-shaped curve of viral count was available for assessment from the 12^th^ of February. Conversely in the other cities besides Beijing, once the PM2.5 levels starting rising, a concomitant rise in viral count was noted. If the viral count curve in Beijing was extrapolated the into early February and late January, highly elevated viral counts correlated with very high levels of PM2.5 The PM2.5 pattern in Beijing in early February and late January initiated with highly elevated mean PM2.5 levels of 155.8µg/m3 (SD+/-73.6) and when the viral counts in Beijing decreased, the PM2.5 also decreased by a margin of 36% to 82.1µg/m3 (SD+/-44.9) (p<0.04).

The origins of COVID-19 are uncertain. There is a possibility that the environmental conditions in the form of atmospheric pollution in China catalysed SARS-CoV-2 genomic evolution from a precursor, possibly a zoonotic predecessor.^13^ Following the initial rise in infection and death rates, human intervention in the form of national and regional lockdown reduced atmospheric pollution. COVID-19 infection and mortality rates abated in some countries only to resurface as a second wave, dominated by the new variant Clade G.

Following the January 2020 lockdown in China, there was a steady decline in airborne particulate matter due to the diminution in the incineration of coal.^13^ Coal consumption in China decreased by approximately 50% following lockdown, from 80 thousand tonnes daily to 40 thousand tonnes per day and consequently was associated with a reduction in atmospheric pollution.^14^ Contemporaneously, the original viral Clade D steadily declined, in parallel with the decrease in atmospheric concentrations of particulate matter. Clade D dominated the epidemic in China from December till mid-February until the persistent variant D614G.^6^ Thereafter Clade G variant slowly dominated the genomic landscape after lockdown beyond mid-February. This may suggest that the changes in airborne particulate matter could have contributed to the transmission and the eventual dominance of Clade G.

Following the emergence of Clade G, the epidemic spread in early March from the Chinese city of Wuhan initially to Qoms in Iran and soon after to Bergamo in Italy. All three cities demonstrated an elevated reproduction number (R_0_) confirming a high transmission rate.^14^ Comparing COVID-19 to the Influenza virus, at its zenith the R_0_ of the H1N1 influenza is 1.4-1.6 while that of COVID-19 was 5.7 at its peak to settle to 3.28.^16,17^ This elevated R_0_ may suggest that there may be another variable accelerating COVID-19 transmission besides human to human transmission.

A characteristic common to the three cities of Wuhan, Qoms and Bergamo, was the elevated atmospheric levels of particulate matter.^15^ Research from the Italian city of Milano detected COVID-19 genes present on particulate matter.^2^ This study showed that compared to control air samples of particulate matter, 34 RNA extractions for the COVID-19 E, N and RdRP genes, detected 20 positive results for one of these genes.^2^

Setti et al (2020) correlated atmospheric levels of particulate matter and the COVID-19 transmission in a number of Italian provinces. The same authors suggested that COVID-19 transmission could be further augmented by particulate matter beyond the social distance of 2 metres up to 10 metres.^18^

When SARS-CoV-2 spread globally, a common pattern was noted whereby the original Clade D was first detected in most cities and when lockdown was established it was slowly displaced by Clade G. This pattern of Clade displacement was demonstrated in all nations except in Iceland. In Iceland, atmospheric pollution levels are perennially low possibly due to the low population density, and the utilization of hydropower and geothermal sources of energy. There was no lockdown variation in atmospheric particulate matter concentrations in Iceland unlike most other countries.^5^

### COVID-19 Viral Clade Counts correlating with PM2.5

Correlations were borne out between the PM2.5 levels and the viral counts. Clade D correlated with PM2.5 levels in Beijing but no correlation was found for the subsequent rise in Clade G. For both Sheffield and Nottingham, both Clades showed correlations between viral counts and PM2.5. Strikingly the pattern of PM2.5 levels were very similar both prior and during the viral count rise: Sheffield 26.9µg/m3 (SD+/-5.9) and 60.3µg/m3 (SD+/-33.3) (p<0.00048); Nottingham 30.3µg/m3 (SD+/− 7.5) and 60.3µg/m3 (SD+/-33.3) (p<0.028).

For Sydney and Cambridge the opposite of Beijing occurred, whereby correlations were found between PM2.5 and viral counts for Clade G only but not for Clade D. Strikingly again, the pattern of PM2.5 levels of both Sydney and Cambridge are very similar both prior and during the viral count rise: Sydney 33.4µg/m3 (SD+/-7.3) and 51µg/m3 (SD+/-27)(p<0.04), and Cambridge 35.4µg/m3 (SD+/-11.1) and 53.8µg/m3 (SD+/-21.5)(p<0.005). These findings may suggest that the levels of PM2.5 may determine the predominant viral Clade.

Clade G may have had a selective advantage in environments with lower PM2.5 levels. Besides elevated levels of atmospheric particulate matter, Clade G may have utilized another source of particulate matter as a vector closer to humans such as tobacco smoking.

### Tobacco Smoking, Pulmonary Pathology and COVID-19

The highest concentration of particulate matter humans are exposed to in a short period of time is during tobacco smoking.^19^ With more than 66% of Chinese males smoking and 70% of the non-smoking population exposed to 2^nd^ hand smoking,^20^ it may well be that the Clade G found a favourable pabulum for COVID-19 transmission and infection through carriage on particulate matter derived from tobacco smoking.^15^ The rate of smoking in adults in the other cities besides Beijing is significantly lower: Cambridge 17.7%, Sheffield 13.9%, Nottingham 20.9%, Sydney 10.5%.^21,22^ This to some extent may explain the significant differential correlations obtained between PM2.5 and both viral Clade counts between all four of these cities.

The adverse effects of particulate matter PM2.5 on respiratory health have been well demonstrated in the scientific literature.^23^ Particulate matter settling in the lung creates free radicals causing oxidative stress on the respiratory epithelium. ^24,25^ Alveolar macrophage cell oxidative stress occurs with exposure to the metallic components of subway PM2.5.^26,27^ Cellular infiltrates follows particulate matter lung exposure resulting in an increase in pulmonary infiltration with neutrophils, eosinophils, T cells and mastocytes.^28,29^ This consequent pulmonary cellular infiltrate can escalate into a cytokine storm which has been implicated in a significant number of SARS-CoV-2 related deaths.^30^

Besides the deleterious effect of particulate matter on anti-microbial respiratory defences and the potential carriage of SARS-CoV-2, there may also be synergism between PM2.5 and the COVID-19 exacerbating lung pathology.^15^ Inhaled particulate matter can result in pulmonary pathology by causing severe inflammatory reaction in lung tissue. Particulate matter appears to influence the renin-angiotensin system which modulates pulmonary inflammatory response. In preclinical studies, lack of angiotensin converting enzyme II was associated with delayed lung repair following PM2.5 induced injury.^31^

The pulmonary port of call for SARS-CoV-2 appears to be the angiotensin converting enzyme II receptor on respiratory epithelial goblet cells.^32^ Tobacco smoking results in an increase in pulmonary goblet cells which in turn increase angiotensin converting enzyme II.^33^ Animal studies have shown that PM2.5 caused vascular oxidative stress in the pulmonary vasculature and heavy metal enhanced protein expression in the pulmonary arterial system.^34^ The finding of COVID-19 genes on particulate matter may suggest that the PM2.5/COVID-19 complex is a potential independent pathological entity. Both components of the PM2.5/COVID-19 complex may act in concert to accelerate pulmonary pathology both in the lung parenchyma and vasculature.^15^

The adverse health effects of particulate matter derived from tobacco smoking and 2^nd^ hand smoking are well established.^35,36^ A study has indicated that USA States with lower restrictions on smoking in public and a higher percentage of the smoking population had a 23% higher incidence of SARS-CoV-2 infection rates than States with more restrictive smoking bans and lower percentage smoking populations.^37^

### Particulate Matter Constituents and the adherence and hydrophobic properties of SARS-CoV-2 Spike Protein

Particulate matter possesses several species differentiated by their elemental composition. The PM species’ components are determined by particulate matter environmental origins.^38^ Similarly particulate matter water content is differentially determined by particulate matter’s main hydroscopic salt components of sodium chloride, ammonium nitrate and ammoniated sulphate.^38^ The ratio of these salts depends on the environmental origin of the particulate matter e.g. particulate matter close to the sea has a higher proportion of sodium chloride. Pulmonary pathology may be exacerbated by the PM predominance of ammonium nitrate and ammoniated sulphate salt components which are certainly more toxic than sodium chloride.^34^ The presence of sodium chloride in the respiratory tract assists in pulmonary anti-microbial defences making bronchial mucus less viscous, encouraging epithelial cleansing ciliary movement.^39^

Particulate matter may have had a bearing on the different geographical distribution between the 1^st^ and 2^nd^ waves of SARS-CoV-2. Coastal cities and islands appear to have been spared the 1^st^ wave in March, only to succumb to the 2^nd^ wave later in August.^40,41^ Coronavirus’ spike protein possesses hydrophobic sites on its N-terminal peptide and the sodium chloride derived-particle based water may have deterred viral adherence to particulate matter.^42,43^ This may not be the case with the 2^nd^ wave whereby Clade G achieved near complete dominance.^7^ The different orientation of Clade G trimeric spike peptides may influence the hydrophobic behaviour of the N-terminal portion of the spike protein peptide.

Particulate matter originating from exhaled tobacco smoke demonstrates a 1.5 times volumetric increase due to the added particle based water attained during its passage through the bronchial tree.^44^ It may well be that if Clade G N-terminal peptide is less hydrophobic than Clade D, that the variant would find particulate matter originating from tobacco smoking a more favourable viral vector. A putative inference is that potentially the asymptomatic SARS-CoV-2 positive smoker may present as a superspreader to both smokers and nearby nonsmokers alike. Moreover the protective effect of masks is lacking during the act of smoking, as obviously face protection has to be removed while smoking, encouraging further COVID-19 transmission.^45^

Other components of particulate matter may also have influenced SARS-CoV-2 adherence to particulate matter. The major component (>60%) of atmospheric particulate is carbon. Subway travel has the highest indoor airborne concentration of particulate matter. Particulate matter found in the environs of subways has a significant haematite component as opposed to the surface carbon-rich particulate matter matter. In the absence of carbon’s adsorbance properties, viral release in the respiratory system may be facilitated by corona-laden haematite rich particulate in subways.^46^

### Limitations of the Study

There was a significant difference in viral counts obtained between cities included in this review. This suggests the strong possibility of variation in the availability of sampling of viral counts between cities. The Beijing data retrieved the latter part of the bell-shaped curve. The assumption that viral counts were higher in Beijing is an extrapolation of the viral count curve. This extrapolation is given greater credence when the curve in Beijing is compared to the other complete curves of the other cities and also correlates with the elevated PM2.5 levels in Beijing in early February and late January 2020. The average daily PM2.5 levels are taken in this study and it must be mentioned that there may be wide variations between the maximum and minimum levels of particulate matter throughout the day including when there may be human exposure to this pollutant. Exposure to atmospheric PM2.5 does not necessarily equate that to humans being similarly exposed to the same levels indoors. Indoor levels may actually be higher if fossil fuels are used for heating purposes. In Wuhan over the past year, natural gas consumption for heating purposes increased 2.8 times this fossil fuel’s utilization by the traditional stove and water heater, potentially producing another indoor source for particulate matter.^47^ The assumption that Clade G utilized particulate matter derived from tobacco smoking is a hypothesis based on the fact that the variant emerged when PM2.5 plummeted in China, combined by the fact that as mentioned earlier more than 66% of Chinese males smoke and 70% of the non-smoking population are exposed to 2^nd^ hand smoking.^20^

## Conclusion

This study suggests that there appears to be an association between PM2.5 levels and COVID-19 infection. The correlations between viral counts and particulate matter may indicate that besides PM2.5’s deleterious effects on pulmonary defences, there may be a viral vector effect that exacerbated the pandemic. Moreover there may be an association with SARS-CoV-2 Clade evolution. The initial Clade D predominated when the PM2.5 were elevated, only to be displaced by Clade G when the atmospheric PM2.5 levels decreased. It may also be inferred that Clade G utilized other sources of this pollutant closer to human respiratory tract, such as particulate matter derived from tobacco smoking employed as a vector for COVID-19 transmission.

## Data Availability

https://www.aqicn.org - waqi.info
https://www.cell.com/cell/fulltext/S0092-8674(20)30820-5

https://www.aqicn.org

http://waqi.info/

